# The Early Food Insecurity Impacts of COVID-19

**DOI:** 10.1101/2020.05.09.20096412

**Authors:** Meredith T. Niles, Farryl Bertmann, Emily H. Belarmino, Thomas Wentworth, Erin Biehl, Roni Neff

## Abstract

**Background:** COVID-19 has disrupted food access and impacted food insecurity, which is associated with numerous adverse individual and public health outcomes.

**Methods:** We conducted a statewide population-level survey in Vermont from March 29-April 12, 2020, during the beginning of a statewide stay-at-home order. We utilized the USDA six-item validated food security module to measure food insecurity before COVID-19 and since COVID-19. We assessed food insecurity prevalence and reported food access challenges, coping strategies, and perceived helpful interventions among food secure, consistently food insecure (pre-and post COVID-19), and newly food insecure (post COVID-19) respondents.

**Results:** Among 3,219 respondents, there was a 33% increase in household food insecurity since COVID-19 (p<0.001), with 35.6% of food insecure households classified as newly food insecure. Respondents experiencing a job loss were more likely to experience food insecurity (OR 3.43; 95% CI, 2.45-4.80). Multiple physical and economic barriers, as well as concerns related to food access during COVID-19, are reported, with respondents experiencing household food insecurity more likely to face access challenges (p<0.001). Significant differences in coping strategies were documented between respondents in newly food insecure vs. consistently insecure households.

**Conclusions:** Since the declaration of the COVID-19 pandemic, there has been a significant increase in food insecurity in Vermont, accompanied by major food access barriers. These findings have important potential impacts on individual health, including mental health and malnutrition, as well as on future healthcare costs. We suggest proactive strategies to address food insecurity during this crisis.

## Introduction

The global COVID-19 pandemic, and social distancing efforts implemented to slow its spread,^1^ have disrupted economies and food systems globally and locally, with extensive food security ramifications. Food insecurity – the lack of consistent physical, social, and economic access to adequate and nutritious food that meets dietary needs and food preferences^2^ – can lead to serious public health consequences. In 2018, 11.1% of American households were considered food insecure at some point in the year, and 4.3% experienced very low food security, characterized by disrupted eating patterns and reduced food intake.^3^ Food insecurity is associated with numerous adverse health outcomes, including chronic conditions such as diabetes mellitus, hypertension, coronary heart disease, depression, and mental health challenges and increased risk of mortality. ^4-7^ Evidence from the United States and Canada has found, on average, health care use ^8,9^ and costs ^4,5,8,9^ to be substantially higher among adults living with food insecurity compared to others.

Food insecurity tracks closely with national and household economic conditions, with trends paralleling unemployment, poverty, and food prices. ^3,10,11^ Given the unprecedented rise in U.S. unemployment since mid-March 2020^12^, models based on data from the 2007-2008 recession predict significant and rapid increases in food insecurity.^13^ Food insecurity is not just a consequence of an inability to afford food, however. The COVID-19 pandemic affects all dimensions of food security, defined by the United Nations to include food availability, accessibility, utilization, and stability.^2^ Food availability has shifted in the short term by consumer panic shopping, but longer-term availability challenges may also unfold. The COVID-19 pandemic threatens accessibility to food through effects on food costs and infrastructure, including changes in food assistance distribution, public transit access, and shortages of certain products. In terms of utilization, market reports indicate widespread changes in food purchasing behaviors. ^14,15^

There are few, if any, peer-reviewed studies using empirical evidence to document actual changes in food insecurity due to COVID-19. Using statewide survey data from Vermont, a U.S. state with a predominantly rural population,^16^ we describe the impact of COVID-19 on household food insecurity among 3,219 Vermont respondents, including their challenges and concerns related to food access, coping strategies, and use of assistance programs, and then discuss public and individual health implications of rising food insecurity.

## Methods

### Survey Development and Recruitment

With feedback from key state-level agencies and hunger relief organizations as well as reviews of relevant literature^3,6^, we developed a survey^17^ to measure food insecurity, food access challenges, and related concerns and experiences. We obtained Institutional Review Board approval from the University of Vermont (IRB protocol 00000873). Using Limesurvey, the instrument was piloted with 25 adults (18 years and older) from the target population. Factor analysis and Cronbach’s alpha on pilot data determined that relevant questions obtained alpha validity above 0.70.^18^ The survey ran online March 29-April 12, 2020. We used four methods for convenience sample recruitment: 1) Paid advertisements via Front Porch Forum, a community-level listserv, which reaches approximately 2/3 of Vermont households^19^; 2) Paid digital ads via Facebook to reach populations under-represented in Front Porch Forum (e.g., males, lower-income households); 3) Listservs of community partners; 4) A University of Vermont press release and subsequent newspaper, radio, and television media.

Vermont’s adult population is 506,631^20^, requiring a sample size of 2,390 to achieve a 95% confidence level for a +/− 2% confidence interval. The survey had 3,953 respondents, including the pilot. Respondents with ZIP Codes outside Vermont (N=59) and empty responses (i.e., people who consented but did not fill in any responses, N=675) were removed, leaving 3,219 eligible responses (Supplementary Figure 1).

Household food security status was determined based on the U.S. Department of Agriculture’s Household Food Security Survey Module: Six-Item Short Form^21^, which was adapted to ask about the time period both “in the year before the coronavirus outbreak” and “since the coronavirus outbreak.” The start of the coronavirus outbreak was set as March 8, 2020, based on the first positive COVID-19 test result in Vermont. According to established scoring procedures from the USDA food security module, respondents classified has having low (2-4 affirmative answers out of six) and very low food security (5-6 affirmative answers) can be combined and referred to as having food insecurity.^21^

### Statistical analysis

To examine differences in household food insecurity during the first weeks of the COVID-19 pandemic, we created three categories of respondents: 1) Households with food security (n=2,282, including households food secure before and since COVID-19 and households who were food insecure at some point in the year before COVID-19 but were no longer food insecure during COVID-19). 2) Households with consistent food insecurity (n=466, both food insecure before COVID-19 and remaining food insecure since COVID-19). 3) Households with new food insecurity (n=258, categorized as food secure before COVID-19, but food insecure since COVID-19).

To determine statistically significant differences between groups, we utilized Kruskal Wallis, Wilcoxon Rank Sum, t-tests, and One-way ANOVAs depending on the distribution of the dependent variable. We utilized a logistical regression model to determine the factors correlated with food insecurity since the COVID-19 pandemic, with coefficients reported in odds ratios. We used all available data to estimate effect sizes and interactions and assumed any missing data were missing at random. Table S1 details the specific questions utilized in this analysis. Future analyses will explore other questions in the survey.

## Results

### Demographic Characteristics of Respondents

Reflecting the demographic composition of Vermont^20,22,23^ the majority identified as non-Hispanic White, lived in rural areas, and had a household income below $75,000 (Table 1, Table S2).

**Table 1.** Characteristics of survey respondent individual and household demographics.

| Characteristic* |  | Respondents (N=3,219) |
| --- | --- | --- |
| Mean age (range) – yr |  | 51.5±15.6 (19-94) |
| Household size (range) – no. |  | 2.7±1.5 (1-12) |
| Gender – no. (%) | Female | 2274 (79.4) |
|  | Male | 539 (18.8) |
|  | Non-binary | 22 (0.8) |
|  | Transgender | 13 (0.5) |
|  | Other (self describe) | 16 (0.6) |
| Race – no. (%) | White | 2669 (96.1) |
|  | Two or more races | 73 (2.6) |
|  | American Indian or Alaska Native | 18 (0.6) |
|  | Asian | 13 (0.5) |
|  | Black or African American | 5 (0.2) |
| Ethnicity – no. (%) | Not Hispanic or Latino | 2783 (98.4) |
|  | Hispanic or Latino | 45 (1.6) |
| Education level – no. (%) | Some high school (no diploma) | 11 (0.4) |
|  | High school graduate (incl. GED) | 260 (9.1) |
|  | Some college (no degree) | 423 (14.8) |
|  | Associates degree / technical school / apprenticeship | 301 (10.5) |
|  | Bachelor's degree | 962 (33.6) |
|  | Postgraduate / professional degree | 910 (31.7) |
| 2019 Household Income – no. (%) | Less than \$12,999 per year | 167 (6.0) |
| | \$13,000- \$24,999 per year, | 332 (11.9) |
| | \$25,000-\$49,999 per year, | 672 (24.0) |
| | \$50,000-\$74,999 per year | 560 (20.0) |
| | \$75,000-\$99,999 per year | 442 (15.8) |
| | \$100,000- \$124,999 per year | 290 (10.4) |
| | \$125,000- \$149,999 per year | 141 (5.0) |
| | More than \$150,000 per year | 193 (6.9) |
| ZIP Code within Census Metropolitan Statistical Area – no. (%) | Yes | 1149 (41.1) |
|  | No | 1649 (58.9) |
| Children in household – no. (%) | Yes | 913 (41.9) |
|  | No | 1267 (58.1) |

\* Plus-minus values are means ±SD. Percentages may not total 100 because of rounding. Percentages are calculated using the number of respondents for that unique question and do not include missing data.

### Food Insecurity Prevalence

We found a one-third increase (33.3%) in food insecurity prevalence (p<0.001) between the year preceding the COVID-19 outbreak, when 18.3% of households (95% CI 16.97%-19.70%) reported experiencing food insecurity at some point, and since the COVID-19 outbreak when the percentage rose to 24.4% (95% CI 22.87%- 25.93%) (Table S3). Among those experiencing food insecurity since the outbreak, 64.4% also experienced food insecurity at some point in the year prior to COVID-19, and were also food insecure since COVID-19, in comparison, 35.6% were newly food insecure. In consistently food insecure households, 57.9% exhibited very low food security since COVID-19 (marked by disrupted eating patterns and reduced intake), while 42.1% had low food security. In newly food insecure households 32.9% exhibited very low food security, while 67.1% had low food security (Table S4) since COVID-19.

Multivariable logit models predicted the factors contributing to higher odds of food insecurity since COVID-19 (Table 2). Respondents experiencing a job loss were significantly more likely to live in a household experiencing food insecurity (OR 3.43; 95% CI, 2.45-4.80), as were those experiencing a furlough (OR 2.73; 95%CI, 1.83-4.06), or a loss of hours (OR 2.05; 95% CI, 1.49-2.82). The odds of experiencing food insecurity since the COVID-19 outbreak were higher among households with children (OR 1.78; 95% CI, 1.36-2.33) while households with higher 2019 incomes had reduced odds (OR 0.55; 95% CI, 0.50-0.61). Finally, women were 1.5 times as likely to experience household food insecurity during COVID-19 compared to men (OR 1.52; 95% CI 1.07-2.16), while increasing age (OR 0.98; 95% CI 0.98-0.99) and a college degree (OR 0.39, 95% CI 0.30-0.50) were associated with reduced odds of household food insecurity.

**Table 2.** Multivariate analysis predicting odds of food insecurity since COVID-19.

| Variable | Odds Ratio | Std. Dev. | P Value | 95% Confidence Interval |  |
| --- | --- | --- | --- | --- | --- |
| Age | 0.98 | 0.00 | 0.001 | 0.976 | 0.994 |
| Race (white) | 0.70 | 0.24 | 0.300 | 0.361 | 1.371 |
| Job Loss | 3.43 | 0.59 | <0.001 | 2.445 | 4.804 |
| Furlough | 2.73 | 0.55 | <0.001 | 1.834 | 4.063 |
| Lost Hours | 2.05 | 0.33 | <0.001 | 1.494 | 2.822 |
| Female | 1.52 | 0.27 | 0.020 | 1.065 | 2.157 |
| Children | 1.78 | 0.24 | <0.001 | 1.362 | 2.329 |
| College | 0.39 | 0.05 | <0.001 | 0.302 | 0.504 |
| Income | 0.55 | 0.03 | <0.001 | 0.502 | 0.608 |
| Urban Metro Area | 0.92 | 0.12 | 0.510 | 0.706 | 1.190 |

### Food Access Challenges and Concerns

Respondents indicated multiple physical and economic barriers to food access during COVID-19, with respondents experiencing household food insecurity significantly more likely to express greater access, availability and utilization challenges than respondents in food secure households (p<0.001) (Figure 1, Table S5). Consistently food insecure households had a higher average prevalence of food access challenges as compared to those in newly food insecure households including trouble affording food (p<0.001), getting food through a food pantry (p=0.004), and knowing where to find help for getting food (p<0.001).

**Figure 1.**
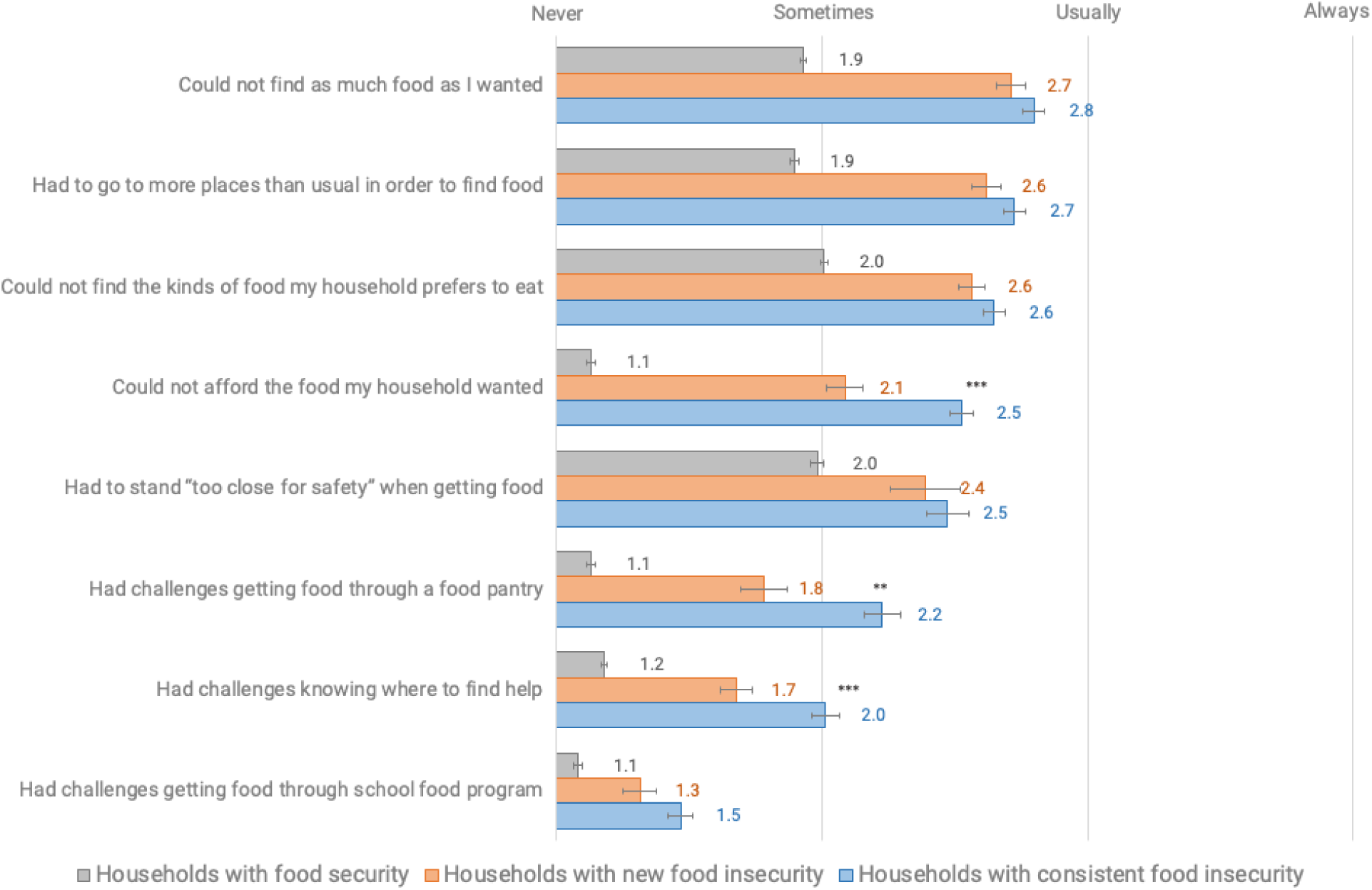
Average frequency of challenging food access situations since COVID-19 among respondents with household food security and food insecurity in a survey of Vermont households, March-April, 2020. (p<0.001 for comparison among all groups). Standard errors shown with brackets. Differences between newly and consistently food insecure shown through stars (***p<0.001), **p<0.01, *p<0.05) and in Table S5.

Respondents experiencing household food insecurity during COVID-19 (both newly and consistently food insecure) were significantly more likely (p<0.001 comparison across all groups) to express higher levels of concern and worry about a variety of potential situations related to food access and COVID-19 (Figure 2, Table S6). As compared to newly food insecure households, consistently food insecure households were also significantly more likely to have higher levels of concern and worry about food access for all situations except for food becoming unsafe (p<0.05, Table S6).

**Figure 2.**
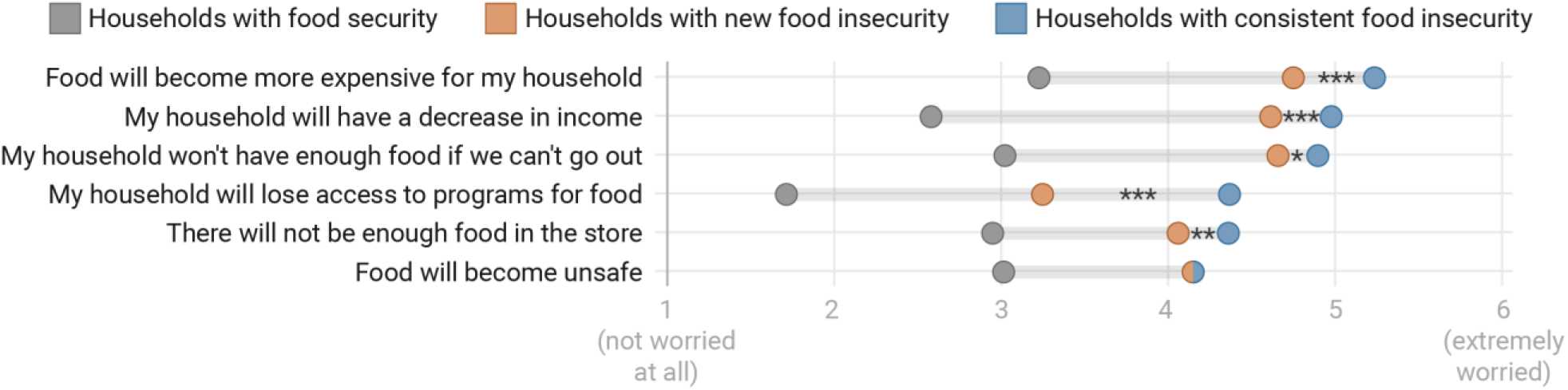
Average level of concern for potential food access situations during COVID-19 among respondents with household food security and food insecurity in a survey of Vermont households, March-April, 2020. (p<0.001 for comparison among all groups). Standard errors shown with brackets. Statistically significant differences were also found between newly and consistently food insecure in all cases except for “food will become unsafe” (shown through stars, ***p<0.001), **p<0.01, *p<0.05) and in Table S6.

### Coping Strategies

Households newly and consistently experiencing food insecurity were significantly more likely (p<0.001) to be implementing coping strategies related to obtaining food as compared to respondents in food secure households, including those related to disrupted eating patterns (i.e. eating less) (Figure 3, Table S7). Consistently food insecure households, as compared to those newly experiencing food insecurity, were also significantly more likely to currently borrow money from friends or family (p=0.01), utilize a food pantry (p<0.001) and utilize government assistance programs (p=0.004), especially the Supplemental Nutrition Assistance Program (SNAP) (p<0.001) (Tables S7 and S8). Households newly and consistently experiencing food insecurity were also significantly more likely (p<0.001 across all group comparisons) to report an intention to implement these same coping strategies in the future for assistance with obtaining food during COVID-19. Among food insecure households, those with consistent food insecurity were more likely to indicate that in the future they would use food pantries (p<0.001), government assistance programs (p<0.001) and to stretch the food they have by eating less (p=0.007) as compared to newly food insecure households (Table S9).

**Figure 3.**
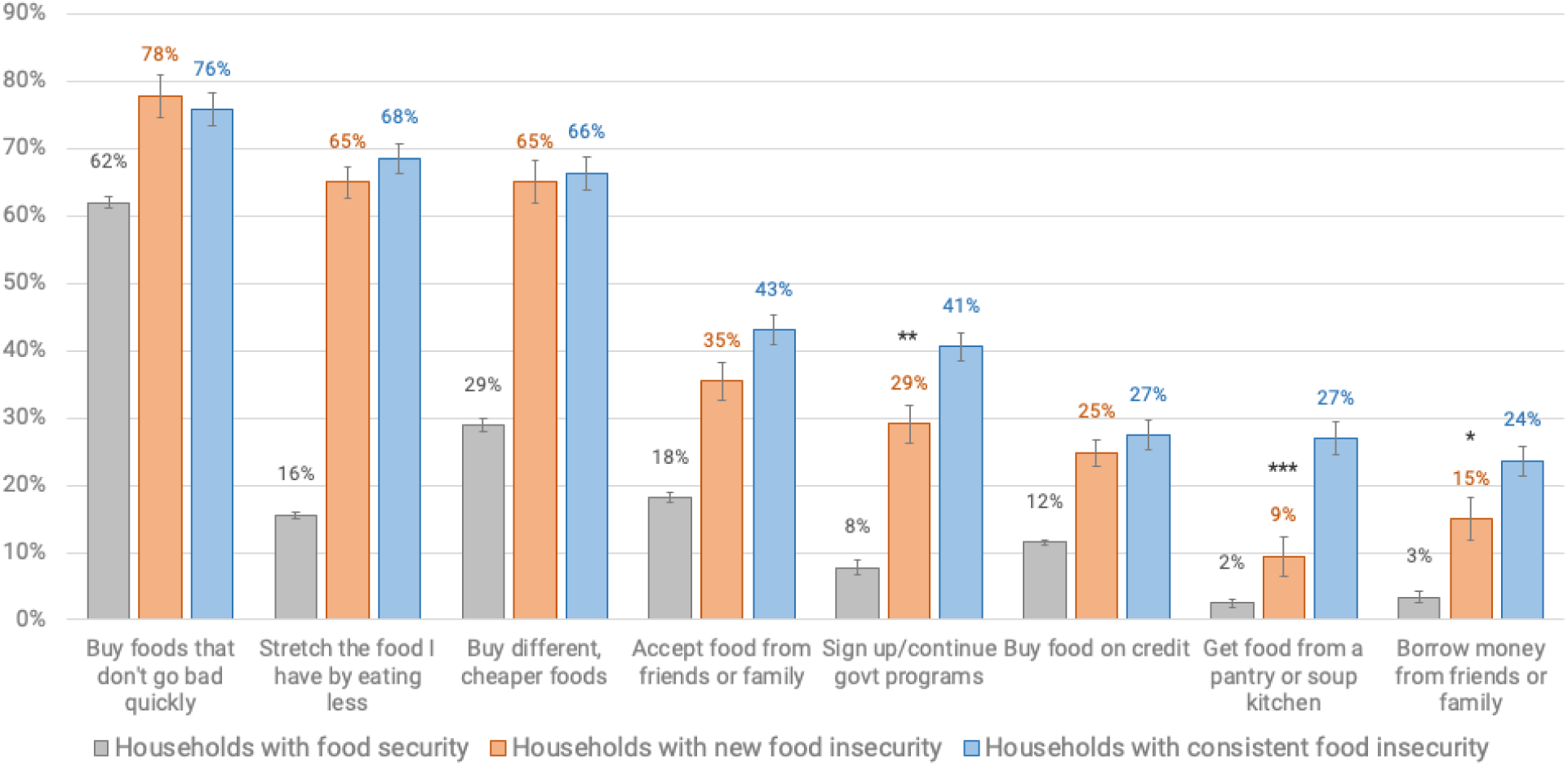
Prevalence of current coping strategies utilized by households with food security and with food insecurity during COVID-19 in a survey of Vermont households, March-April, 2020. (statistical differences among all groups p<0.001). Statistical differences between newly and consistently food insecure shown through stars (***p<0.001); **p<0.01, *p<0.05) and in Table S7.

### Desired Interventions

Compared to food secure households, new and consistently food insecure households were significantly more likely (p<0.001) to find strategies to address physical or economic food access challenges helpful during COVID-19 (Table S10). Consistently food insecure households were also significantly more likely than those in newly food insecure households to find access to public transit, extra money for food or bills, increased benefits of food assistance programs, information about food assistance programs (all p<0.001), help with administrative food assistance problems (p=0.001), and support for food delivery costs (p=0.033) more helpful (Table S10).

## Discussion

This statewide survey in Vermont documented a statistically significant increase in food insecurity since the state’s first reported case of COVID-19 and the stay-at-home executive order. We demonstrate a one-third increase in household food insecurity among respondents, with individuals experiencing job loss or disruption at significantly greater odds of experiencing household food insecurity since COVID-19 as compared to other demographic controls. Further, we find that the majority of consistently food insecure households and one third of newly food insecure households were classified as having very low food security, marked by disrupted eating and cutting meals or going hungry. Fully two-thirds of Vermont respondent households with food insecurity during COVID-19 are already eating less to stretch their food. The findings indicate challenges to all food security dimensions: including economic and physical access, availability, utilization, and stability, and may have profound potential health impacts.

We further demonstrate physical and economic barriers to food access during COVID-19 and the coping strategies of respondents in food insecure households. Previous research^10,11^ suggests links between job loss and food insecurity, indicating that the profound increase in Americans experiencing job loss and disruption^24^ will present acute and large-scale impacts across the population. Since Vermont unemployment claims reflect the national trend, these results likely reflect a broader U.S. phenomenon of rising food insecurity rates, evidenced by early non-peer reviewed studies.^25,26^ In addition to these new economic barriers, the pandemic presents many new physical barriers for food access with reductions in public transportation and new distribution models, and in a rural state like Vermont, a lack of income for transportation costs including fuel. In rural areas where food assistance programs such as food pantries are limited, closures due to illness, social distancing, or lack of volunteers may be particularly challenging. This presents opportunities to expand food pantries, support mobile pantry units, as well as encourage the expansion of programs such as fruit and vegetable prescription programs, shown to positively affect food security^27^ and improve health outcomes^28^. Ultimately, this research demonstrates a need to increase food assistance programs and provide resources to remove food access barriers now, and likely in the future, during state and national economic and health emergencies.

This rise in food insecurity presents many potential health impacts. Food insecurity is negatively associated with health outcomes^5,6^ and some evidence indicates it is positively associated with poor diet quality. ^29,30^ Further, higher rates of anxiety and mental health disorders among both children and adults have been documented in food insecure households.^6,29^ Indeed, survey respondents in this study experiencing household food insecurity demonstrated significantly higher rates of concern and worry about food. Disrupted eating, found in two-thirds of respondent households with food insecurity, is associated with decreased immune function and negatively impact mental and emotional health^29^. Further research is needed to understand how food insecurity during the COVID-19 pandemic relates to diet quality, particularly if disrupted eating patterns persist and increase.

Healthcare providers can address food insecurity through simple interventions. Screening for food insecurity and providing resources now may reduce short and long-term consequences including the potential long-term impacts on child health outcomes associated with the duration of household food insecurity ^31^ and higher health care expenditures associated with food insecurity^9^. The Hunger Vital Sign, a validated two-question food insecurity screening tool based on the USDA Household Food Security Survey Module^32^, can quickly determine risk for food insecurity in clinical and community settings. This tool is widely utilized, especially in pediatrics^33,34^, and could be made standard in health care and other service settings during COVID-19 and beyond. Providers could refer families in need to locally available resources or to United Way, which aggregates these resources locally. However, during this heightened time of unemployment, there is also potential for government agencies, particularly those distributing unemployment benefits, to help connect families in need to available resources as well.

Importantly, this research demonstrates there are still a significant number of food insecure households which, even if aware of food assistance programs, may not utilize them. Low rates of seeking assistance in our results, especially among newly food insecure households, may be partly related to the stigma associated with assistance programs. ^35,36^ Prior research suggests that populations living outside major metropolitan areas may be more likely to utilize friends and family for support^37^ and to see government assistance programs as a “last resort”.^38^ However, with social distancing and widespread financial challenges, such personal safety nets may be eroded, and these households may be particularly vulnerable. Additional research is needed to understand barriers to utilize food assistance programs, especially among those that may be newly food insecure since COVID-19.

This study suggests the first population-level impacts of COVID-19 and social distancing on food insecurity. The limitations are partly rooted in the need to rapidly administer this survey in the early days of the pandemic, to provide data that can be tracked over time. Though our respondent population matches statewide census statistics closely on many metrics, this was a convenience sample; further research is expanding these results using similar questions with representative samples across states and populations. Further, we utilize an internet-based survey given the necessity of social distancing during COVID-19 and the need for a rapid response, which may limit the capacity of some people to participate, though 81% of Vermonters do have internet plans^23^. The study’s strengths include its large sample size, early administration, population-based assessment, and survey instrument addressing the multiple dimensions of food security.

We implemented this survey in the beginning of a stay at home order and COVID-19 economic impacts. As such, it is likely that many respondents experiencing job loss or disruption had not yet received unemployment benefits and federal stimulus checks were not distributed. Future research will examine the evolution of food security impacts, and how various interventions, including the CARE Act and unemployment benefits, as well as food assistance expansion and health care screenings, may affect food insecurity outcomes as COVID-19 unfolds.

## Data Availability

Data will be made available upon publication

